# CRISPR-cas13 enzymology rapidly detects SARS-CoV-2 fragments in a clinical setting

**DOI:** 10.1101/2020.12.17.20228593

**Authors:** Wahab A. Khan, Rachael E. Barney, Gregory J. Tsongalis

**Affiliations:** Department of Pathology and Laboratory Medicine, The Audrey and Theodore Geisel School of Medicine at Dartmouth College, Hanover, NH 03755, USA; Laboratory for Clinical Genomics and Advanced Technology, Department of Pathology and Laboratory Medicine, Dartmouth Hitchcock Medical Center, 1 Medical Center Drive, Lebanon, NH 03756, USA

## Abstract

The well-recognized genome editing ability of the CRISPR-Cas system has triggered significant advances in CRISPR diagnostics. This has prompted an interest in developing new biosensing applications for nucleic acid detection. Recently, such applications have been engineered for detection of SARS-CoV-2. Increased demand for testing and consumables of RT-qPCR assays has led to the use of alternate testing options in some cases. Here we evaluate the accuracy and performance of a novel fluorescence based assay that received EUA authorization from the FDA for detecting SARS-CoV-2 in clinical samples. The Specific High-Sensitivity Enzymatic Reporter UnLOCKing (SHERLOCK) technology forms the basis of the Sherlock CRISPR SARS-CoV-2 kit using the CRISPR-Cas13a system. Our experimental strategy included selection of COVID-19 patient samples from previously validated RT-qPCR assays. Positive samples were selected based on a broad range of cycle thresholds. A total of 50 COVID-19 patient samples were correctly diagnosed with 100% accuracy (relative fluorescence ratios: N gene 95% CI 23.2-36.3, ORF1ab gene 95% CI 27.6-45.4). All controls, including RNase P, showed expected findings. Overall ratios were robustly distinct between positive and negative cases relative to the pre-established 5-fold change in fluorescence read output. We have evaluated the accuracy of detecting conserved targets of SARS-CoV-2 across a range of viral loads using the SHERLOCK CRISPR collateral detection reaction in a clinical setting. These findings demonstrate encouraging results, especially at a time when COVID-19 clinical diagnosis is in high demand; often with limited resources. This approach highlights new thinking in infectious disease identification and can be expanded to measure nucleic acids in other clinical isolates.

## Introduction

The novel infectious severe acute respiratory syndrome coronavirus 2 (SARS-Cov-2) emerged in Wuhan, China in late 2019 and has since become a global pandemic^1^. From reports of the initial set of infected patients^2–4^, emerging evidence continues to reflect that some individuals experienced severe conditions requiring intensive care and hospitalization with a significant number succumbing to the virus. In the majority of patients, the clinical course typically ranged from marked fever, myalgia, pneumonia to dyspnea and respiratory distress^2–4^. As we learned more about the disease course, it became apparent that most individuals present with mild symptoms, but more importantly, asymptomatic or low-level infected individuals can become carriers who further potentiate the spread of the COVID-19 disease ^5,6^.

Mitigating the spread of the virus clearly involves accurate, efficient and increased testing with a simultaneous public health surveillance effort. The gold standard test uses quantitative reverse transcriptase polymerase chain reaction (RT-PCR) approach. RT-PCR based approaches for SARS-CoV-2 typically require detection of multiple targets. For this, several pre-requisites need to be in place such as having hydrolysis sequence-specific probes that need to be labeled with a distinct fluorescent dye and an appropriate quencher moiety. The emission maxima of the dyes must not overlap with each other and reactions need to be carried out on an appropriate real-time cycler that supports multiplex analysis. Further, for downstream quantification, standard curves are necessary for analysis and the efficiency of the method may vary between runs and reactions. Taken together, although PCR is very effective in optimized scenarios, these pre-requisites can limit the use of PCR testing in locations outside central laboratories. Therefore, frequent, rapid, and portable testing is necessary; especially with the already limited resources for RT-PCR based reagents and consumables. This has prompted new strategies for viral RNA detection of SARS-CoV-2. Clustered regularly interspaced short palindromic repeats (CRISPR) can complement RT-PCR testing and provide comparable sensitivity and high specificity. This approach uses guide RNA enzymes from the Cas family (i.e. Cas12, Cas13) that are engineered with a programmable spacer sequence and nucleotide binding domains that can cleave the nucleic acid target of interest^7,8^.

In the current study, we employ the <u>S</u>pecific <u>H</u>igh Sensitivity <u>E</u>nzymatic <u>R</u>eporter Un<u>LOCK</u>ing (SHERLOCK) technology that was initially described as a means to rapidly detect nuclei acids in human health applications. In particular, this was applied to instances where different strains of the Zika and Dengue viruses as well as other pathogenic bacteria needed to be distinguished ^9,10^ It has since been used to identify mutations in cell-free tumor DNA in the background of genomic DNA and has broad applications in genotyping human DNA^9^. The current approach naturally leverages SHERLOCK as a CRISPR-based diagnostic platform to detect SARS-CoV-2 viral RNA. The main principal behind SHERLOCK is that in the presence of SARS-CoV-2 viral RNA, a Cas13 enzyme complexed with a virus targeting RNA will be activated and will cleave the viral RNA resulting in a collateral RNase activity^7,11^.

The sensitivity of the assay to detect synthetic SARS-CoV-2 viral RNA was determined by Sherlock Biosciences as part of its emergency use authorization (EUA) and was successfully submitted to the FDA (https://www.fda.gov/medical-devices/coronavirus-covid-19-and-medical-devices/sars-cov-2-reference-panel-comparative-data). Each viral RNA target (N, O) was tested in triplicate over a range of 10 dilutions. The estimated limit of detection for the N and O target was determined to be 0.9 and 4.5 copies/µL of viral transport media (VTM). The analytical sensitivity was confirmed by testing at 1.5x the estimated limit of detection, which was determined to be 6.75 copies/µL of VTM from 19 out of 20 samples with a 95% detection rate (https://www.fda.gov/media/137747/download). Here we extend this work and present a clinical evaluation of the SHERLOCK approach in a CLIA-certified molecular diagnostics laboratory performing high complexity testing for detection of SARS-CoV-2.

## Results

### Clinical specimen testing

For our SHERLOCK experimental design, we utilized anonymized clinical patient samples previously tested using the SARS-CoV-2 CDC assay on the Applied Biosystems 7500Fast Dx instrument implemented near the start of the pandemic by our molecular laboratory ^12^. These patient sample results were also subsequently confirmed using orthogonal technologies from Abbott (m2000 real-time system) and ChromaCode (HDPCR™ SARS-CoV-2 real-time PCR assay). Samples within three ranges of cycle threshold (Ct) values (low positive: 30-36; mid positive: 15-30; high positive: 5-15) were included in the analysis where the Ct value is inversely proportional to the viral copy number. A total of 33 positive (SARS-CoV-2 detected) and 17 negative samples were analyzed (n = 50 total) using the SHERLOCK Cas13 technology.

### SHERLOCK assay performance

The SHERLOCK assay correctly confirmed all 50 previous clinical sample results with 100% sensitivity (**Table 1, Figure 1**) and 100% positive-negative predicative agreement (Table 2). A series of pooled positive samples that were tested in triplicate also showed consistent results and reproducibility of the assay. The raw signal output graph on the BioTek Gen5 software clearly demarcated a positive result relative to a negative one with a marked increase in relative fluorescence (**Figure 2**). Ratios calculated from the raw fluorescence values at t_10min_ for test samples were nearly 45 to 55 fold higher for the Nucleocapsid (“N”) and Open Reading Frame (*ORF1ab*, “O”) gene targets (ratios > 5) respectively for a positive Cas13 detection relative to a negative finding. This further suggests that the collateral cleavage activity of the CRISPR complex is robust at differentiating infected from non-infected patients in our cohort with high accuracy. All positive, negative and internal template controls, RNase P *POP7* gene (“RP”), performed as expected with normalized fluorescence ratios well below a 3-fold increase cut-off (**Table 1**).

**Table 1.**
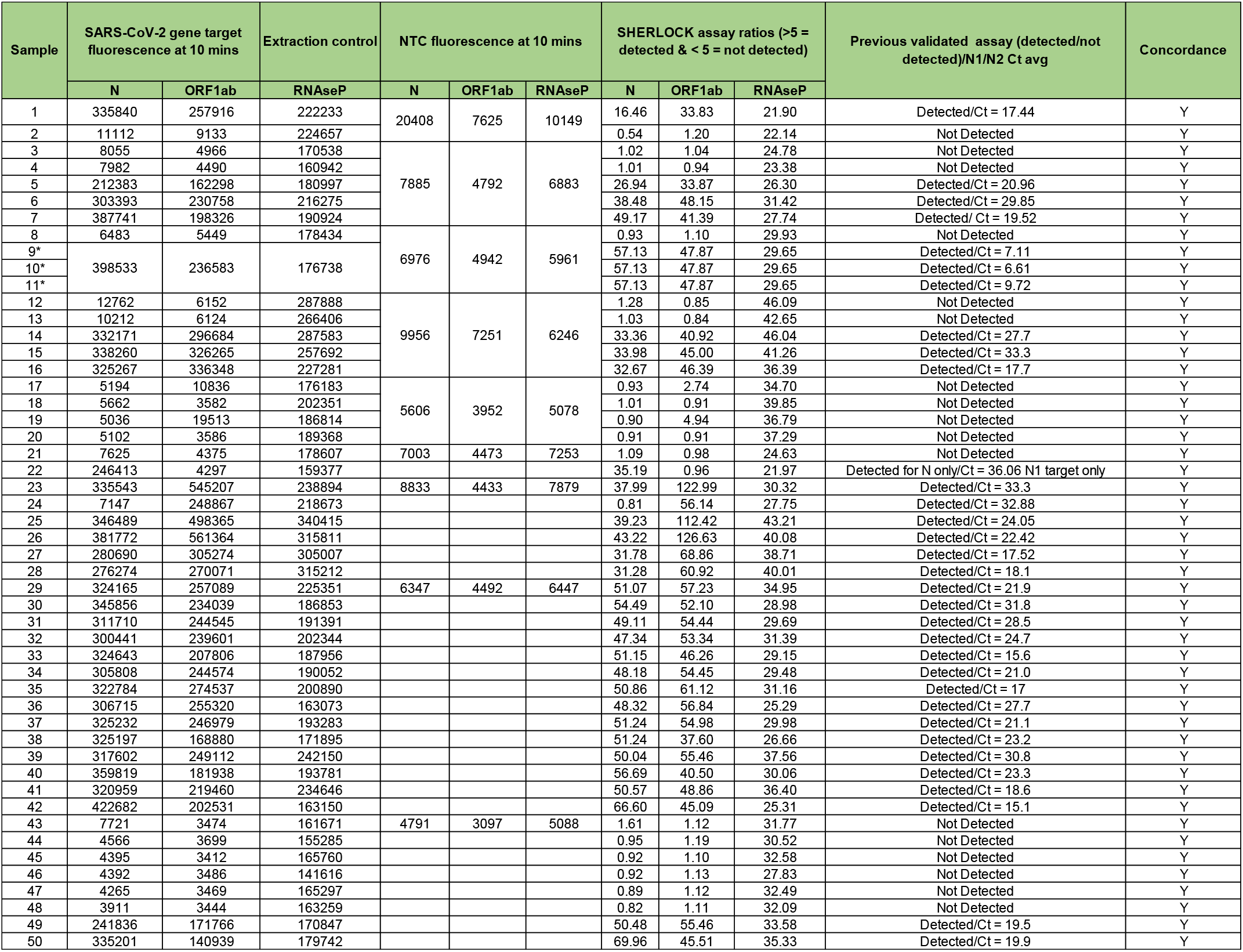
Summary of clinical SARS-CoV-2 nasopharyngeal samples processed on the SHERLOCK CRISPR-Cas13 assay with comparable concordance to the CDC assay. * Indicates pooled set of samples.

**Table 2.**
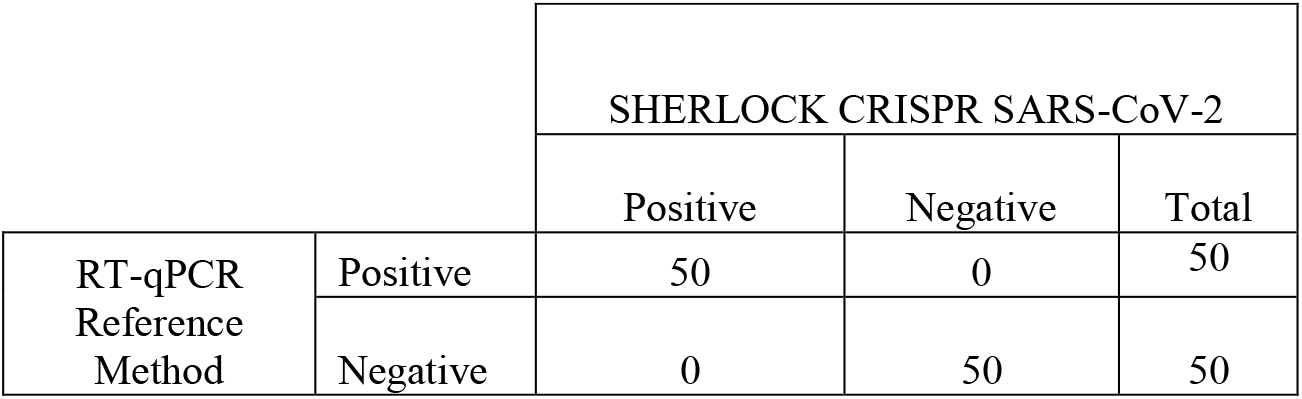
Summary of RT-qPCR results compared to Sherlock SARS-CoV-2 kit detecting positive and negative nasopharyngeal clinical samples (n =50). No false positive or negative results were noted and the sensitivity/specificity along with positive-negative predicative value was at 100% (N gene: 95% CI 23.2 to 36.3; ORF1ab gene: 95% CI 27.6 to 45.4).

|  |  | SHERLOCK CRISPR SARS-CoV-2 |  |  |
| --- | --- | --- | --- | --- |
|  |  | Positive | Negative | Total |
| RT-qPCR<br>Reference<br>Method | Positive | 50 | 0 | 50 |
|  | Negative | 0 | 50 | 50 |

**Figure 1.**
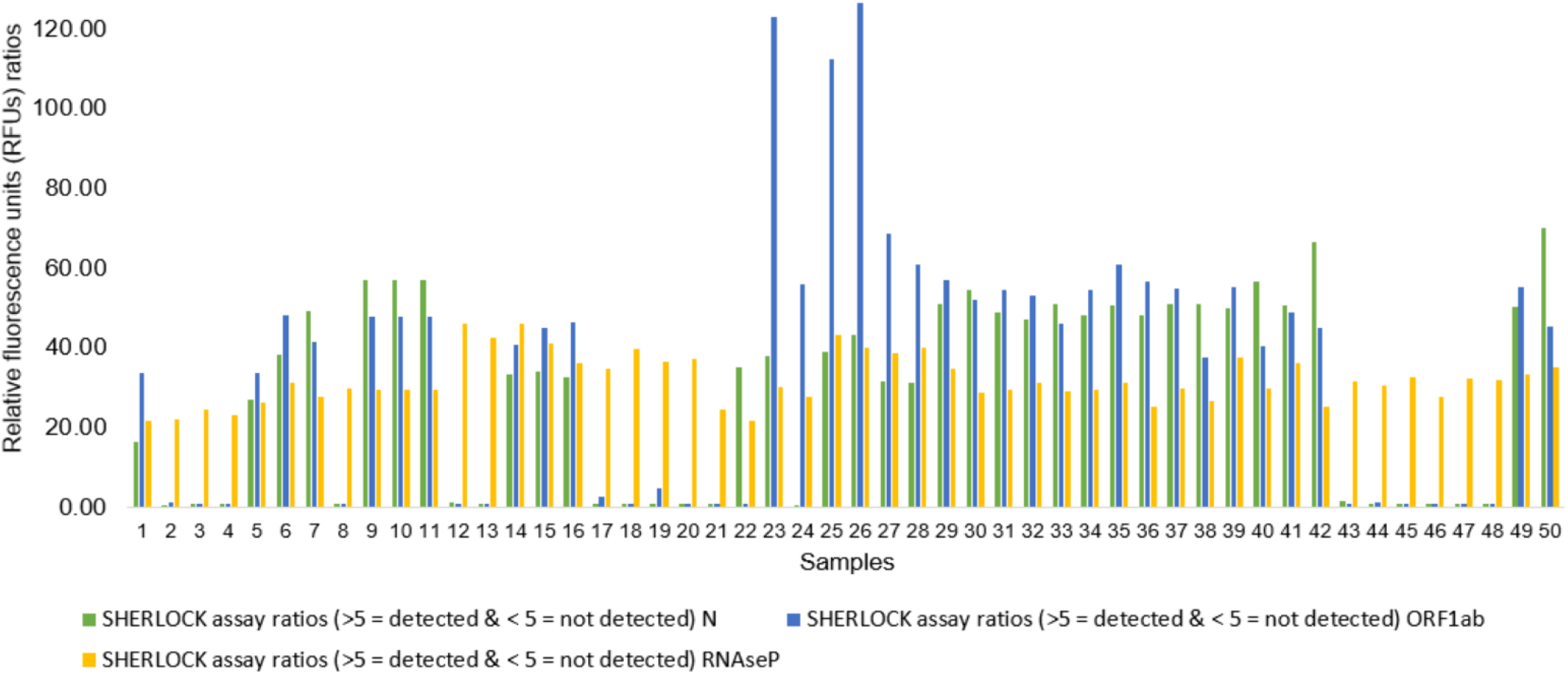
Reaction fluorescence ratios across a range of SARS-CoV-2 samples detected with the SHERLOCK CRISPR-Cas13 system. The Y axis depicts the relative fluorescence units (RFU) calculated from the raw fluorescence detected for each sample (X-axis) normalized to the no template control. The final values spanning over a 10-minute interval were used in these calculations. In scenarios where RFU from the N or ORF1ab SARS-CoV-2 gene target is not detected, the RNAseP extraction control was consistently present (ratio > 5) in each clinical sample. Negative specimens are marked by an absence of a discernable green or blue bar (RFU ratio < 5), where as a positive specimen revealed a prominent signal well above a RFU ratio of 5.

**Figure 2.**
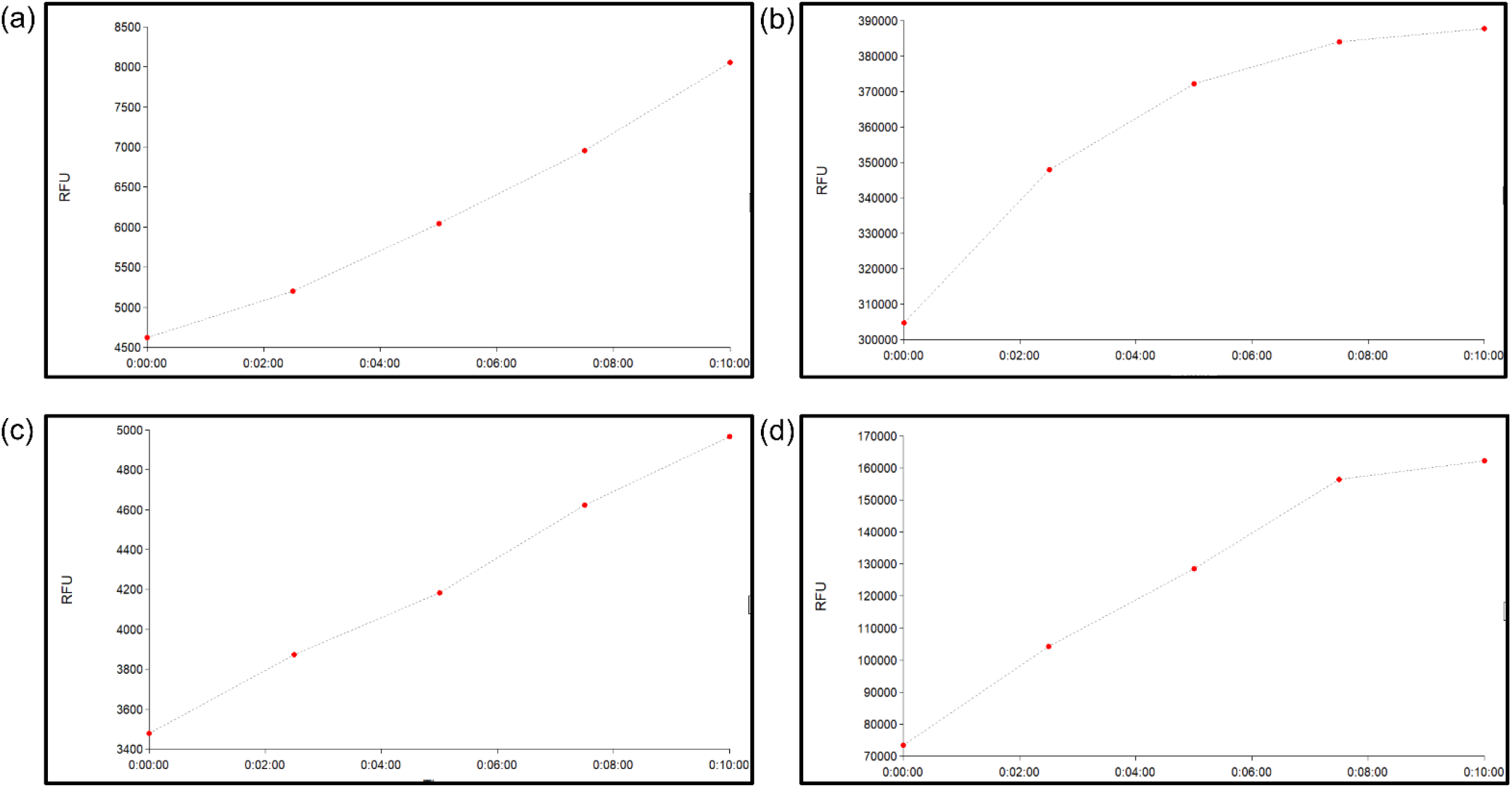
Raw fluorescence (Y-axis) shown as a function of time (X-axis in minutes) from representative samples processed on a standard multimode plate reader. (a) N – gene target not detected in this clinical specimen whereas (b) a prominent spike in fluorescence is noted from a separate clinical sample in which the N – gene target is robustly detected. A similar pattern is shown in the bottom two panels where the *ORF1ab* target is not detected (panel c) versus detected (panel d). Of note, the Y-axis values are an order of magnitude higher between the two scenarios (e.g. detected/not detected) due to the collateral activity of the CRISPR Cas13 guide RNAs to cleave and release fluorescence in the presence of SARS-CoV-2.

### Potential for scale-up

In terms of sample set-up and processing, we have currently observed optimal detection in a 384 well plate system, given the volumes of the Cas13 detection reaction used as part of the experimental design. This small diameter of the sample vessels in which the CRISPR reactions commence maximally retains fluorescence and promotes favorable molecular collisions during the SHERLOCK step. However, increasing the Cas detection volume may allow for comparable detection using a 96-well format. We can accommodate approximately 200 samples on one plate if every other well in a 384-well column configuration is omitted to pre-emptively minimize any risk for contamination. On a Hamilton STAR system or similar ^13^, both a 384 and a 96 probe head with rocket tips can potentially process up to 5 plates for an output of approximately 1000 samples in an eight-hour shift.

## Discussion

We have successfully demonstrated the diagnostic potential of SHERLOCK CRISPR-Cas13 enzymology for the detection of SARS-CoV-2 in 50 previously tested patient samples. In this proof of principal study, all samples subjected to this approach were in keeping with results reported for detecting SARS-CoV-2. A major advantage of CRISPR based diagnostics is that it is highly specific, because the Cas13a RNase remains dormant until it binds to its specific programmed RNA target ^14^. In addition, this assay has the potential to be scaled up for higher throughput testing capacity.

CRISPR-Cas13 as well as Cas12 have also shown efficacy in detecting other viral diseases besides COVID-19 ^15^. Similarly, SHERLOCK methods have exploited CRISPR-mediated detection of SARS-CoV-2 with comparable sensitivity to RT-PCR^16^. These tests, and earlier versions of this technology, have rapidly detected nuclei acids in human health applications^10^. The SHERLOCK approach, in particular, has been compatible not only with fluorescence read outputs but also lateral-flow assays^10,16^. Moreover, flexibility in constructing crRNAs and other RNA sequences continue to push the detection limit and turnaround time for CRISPR diagnostics. Of note, a limit of detection of 12 copies/µL and a 45 minute run time has been reported for SARS-CoV-2 RNA detection using the Cas12 system^17^. This assay, termed DETECTR was given EUA status, and examines the N2 portion of the N gene. The Sherlock CRISPR SARS-CoV-2 kit corroborates these findings and expands the repertoire of viral genomic analyses to the Cas13 CRISPR-Cas effector family using fluorescence-based detection. By contrast to the DETECTR assay, the SHERLOCK approach used in the current study examined two SARS-CoV-2 gene targets (N and ORF1ab). Overall Sherlock’s CRISPR SARS-CoV-2 kit exhibited a limit of detection down to 6.75 copies/µL, as mentioned above, and had an approximate process time of 50 minutes.

As advances in COVID-19 diagnostics emerge, CRISPR testing, because of its many advantages, will be at the forefront. ^11,14^. This is further underscored by the 2020 Nobel prize recently awarded towards this disruptive technology that has made scientific history^18,19^. In the context of the current work, SHERLOCK technology is also poised to rapidly detect SARS-CoV-2 RNA from other sample matrices.

The COVID-19 pandemic has accelerated CRISPR diagnostics and we are only at the beginning of this revolution. One of the unique features of the SHERLOCK approach applied here is that viral RNA can be directly detected with high accuracy and the CRISPR-Cas13 enzyme is rapidly programmable to detect any RNA based target. From a public health and governance perspective, the development of fast, accurate and a portable point of care diagnostic tests is critical for imminent infectious diseases outbreaks and surveillance^20,21^. A test such as the one deployed in our clinical setting will be indispensable in the current pandemic as it does not require large equipment and obviates the need for in-demand PCR consumables.

## Methods

### Overview of experimental protocol

The Sherlock CRISPR SARS-CoV-2 kit has two main steps and was completed in under 2 hours from a brief reverse transcriptase loop-mediated amplification (RT-LAMP)^**22**^ step to direct CRISPR detection of the RNA targets in 10 minutes. In the first step, SARS-CoV-2 RNA is reverse transcribed to DNA and the DNA is amplified by a strand displacing DNA polymerase. The second step initiates transcription of the DNA and activates collateral cleavage activity of CRISPR complex programmed to the target RNA sequence (i.e. programmable crRNA)^**11**^. As Cas13 cleaves the reporter RNA molecules, separating the two ends, a fluorescent signal is detected on a standard plate reader (**Figure 3**). SHERLOCK technology was designed to detect fragments of the N and the O genes of SARS-CoV-2. A third target, the human RP gene, acted as an internal extraction control. Of note, and unlike most molecular assays, SHERLOCK identifies SARS-CoV-2 RNA rather than cDNA providing increased target specificity. In the current experimental protocol, upper respiratory specimens mainly from nasopharyngeal swabs were tested.

**Figure 3.**
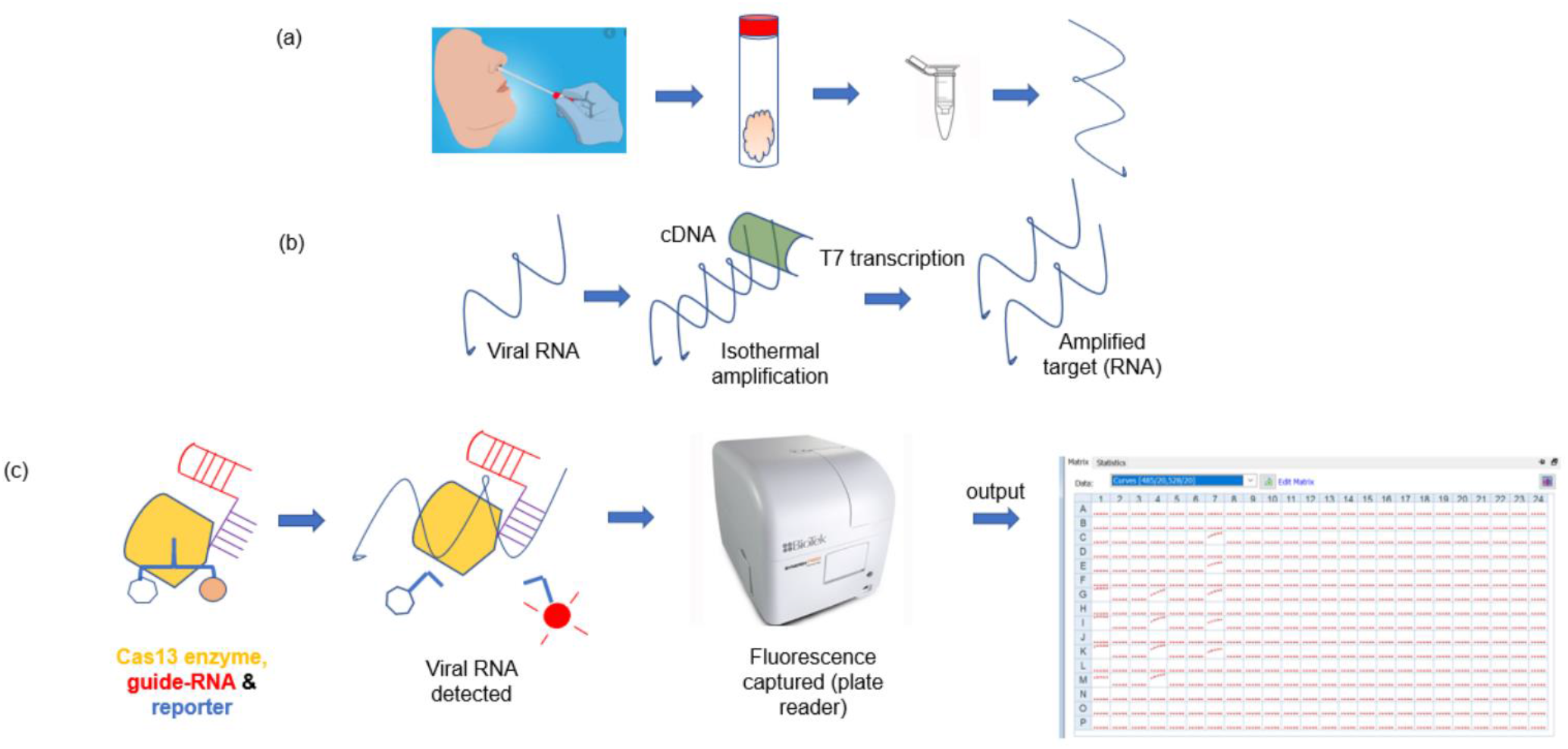
Basic workflow of CRISPR-Cas13 detection of SARS-CoV-2 as it relates to pre-processing, analytical and post-analytical steps. (a) Upon collection of nasopharyngeal swab and nucleic acid extraction, the purified RNA is subjected to an (b) isothermal amplification (green depicts the reverse transcriptase enzyme). (c) The amplified target upon being bound by the Cas13 enzyme (yellow), with its guide RNA (purple-red) cleaves the reporter (blue). A standard output generated on a plate reader is shown form which data export into an excel format is performed. All sequence of events are shown in succession left to right in panels a-c.

### Specimen collection and preparation

All sample collection and preparation steps were performed in accordance with CDC interim guidelines (https://www.cdc.gov/coronavirus/2019-ncov/lab/). The extraction of RNA from nasopharyngeal swabs placed into VTM was carried out using the QIAGEN EZ1 virus mini v 2.0 kit on the EZ1 advanced XL instrument. Samples were eluted in a final volume of 120µL. A total of 8 µL of the extracted RNA was used in a given CRISPR reaction and the remaining stored in aliquots at −80°C for subsequent use. The RT-LAMP master mix (IDT, catalog # 10006968) was prepared separately with primer pairs (10x concentrate) targeting the N, O, and the RP gene as an internal quality control step. A no template or NTC control along with a SARS-CoV-2 positive control (BEI; Catalog # NR-52285) was also included in final reaction volumes of 20µL, the latter diluted to a stock concentration of 4800 copies/µL.

### CRISPR Cas preparation and detection

The Cas master mix reaction with the “N” “O” and “RP” specific guide RNAs (crRNA), reporter reagent, and LwaCas13a enzyme was prepared according to manufacturer’s instructions (IDT, Sherlock™ CRISPR SARS-CoV-2 kit). This mixture was distributed into separate reaction strip tubes of 20 µL aliquots. To each respective 20 µL Cas13 mix, 5 uL of RT-LAMP reaction containing the test sample and appropriate controls was added and the reaction mixture vortexed for ∼10 seconds. A final volume of 20 µL of the mixture was transferred, based on a pre-designated layout indicating the spot location of each sample, to a 384-well flat low-volume clear bottom assay plate (Corning, catalog # 3540). This step was performed in an air clean system to avoid cross-contaminants. The plate was protected with a microamp optical adhesive film (Applied Biosystems, catalog # 4311971), prior to loading onto a pre-warmed multimode plate reader (BioTek Synergy NEO2). All steps was performed in a unidirectional workflow.

### Data processing and analysis

The specific plate reader configuration settings for initiating the analysis are provided in **Figure 4**. Following the 10 minute sample analysis protocol, raw fluorescence data from each well of the assay plate was transferred into an excel document. The raw data were then normalized by dividing target reaction fluorescence accumulated at time point 10 minutes (t_10 min_) of the test sample to that of the NTC reaction, also measured at the same time point interval. This generated a quantitative result in the form of a ratio. Briefly a result was considered “detected” if a given SARS-CoV-2 target reaction produced a fluorescence ratio that was greater than or equal to a 5-fold increase in florescence measured at 10 minutes for the test sample over the corresponding NTC reaction control. And if this ratio calculated less than a 5-fold increase in reaction fluorescence, the result was considered as “not detected” for SARS-CoV-2. Additional specifics to the ratio calculation and interpretative outputs are tabulated in **Table 3**.

**Table 3.** List of ratio combinations and interpretation for reporting SARS-CoV-2 results on clinical samples. Three targets are listed, including the RNAse P *POP7* gene which serves as a control for the extraction of the clinical sample in the absence of a positive SARS-CoV-2 result. Note the experiment should also be considered invalid if the NTC (not shown in table) shows a ratio >5.

| N | ORF1ab | RNase P | Interpretation |
| --- | --- | --- | --- |
| $\geq 5$ | $\geq 5$ | n/a | Detected |
| $\geq 5$ | $\leq 5$ | n/a | Detected |
| $\leq 5$ | $\geq 5$ | n/a | Detected |
| $\leq 5$ | $\leq 5$ | $\geq 5$ | Not Detected |
| $\leq 5$ | $\leq 5$ | $\leq 5$ | Invalid |

**Figure 4.**
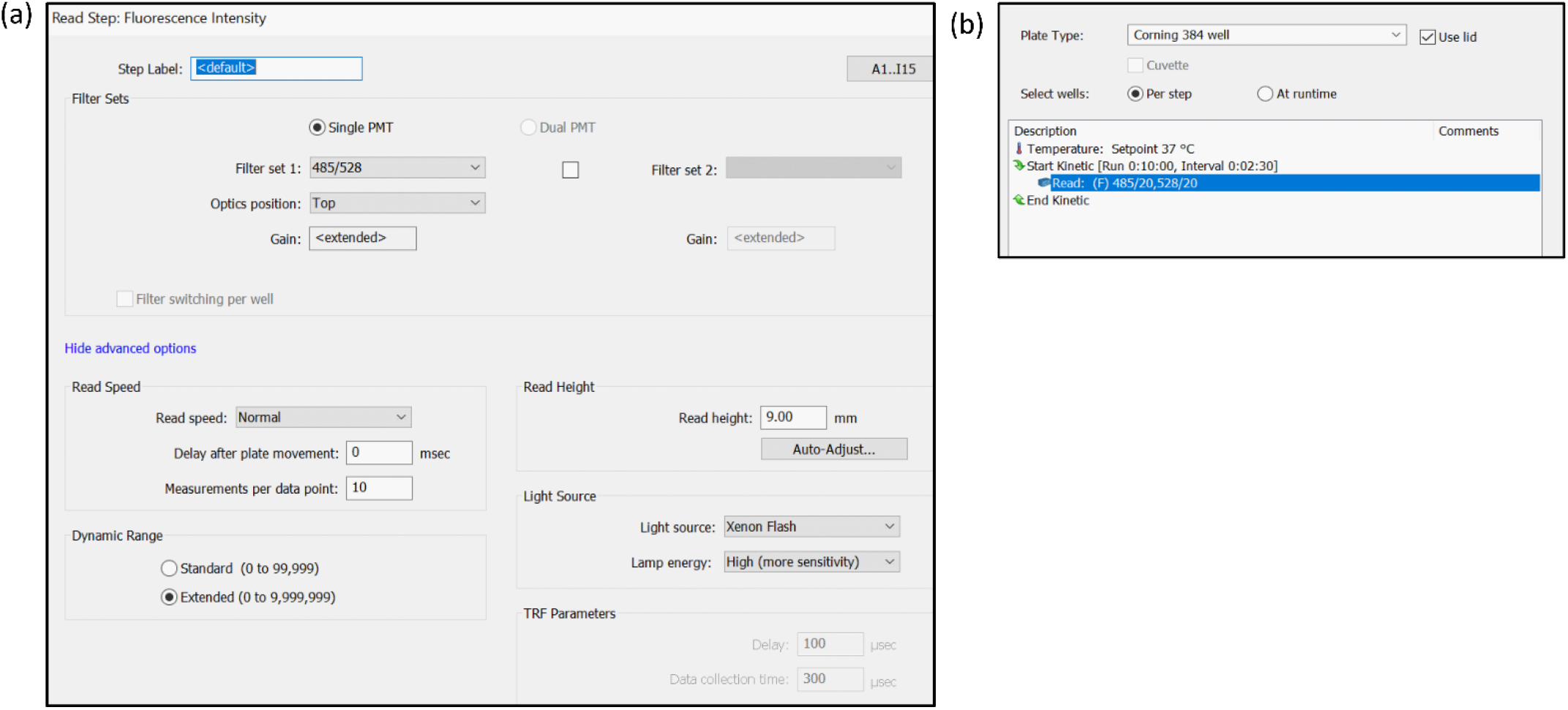
Settings on BioTek plate reader applied during SARS-CoV-2 detection step. (a) The read speed was set to normal and standard filter excitation and emission spectra were all collected at 485/528 respectively. (b) Settings for collecting data is broken up into 2.5-minute intervals with a total kinetic read time of 10 minutes.

## Data Availability

Correspondence and requests for material should be addressed to the last author on the manuscript

## Acknowledgements

We thank members of SHERLOCK Biosciences for sharing reagents, initial EUA data, and valuable feedback for this project. We thank Yvonne FitzGerald (BioTek Instruments Inc., part of Agilent) for technical support on the plate reader. The authors acknowledge the support of the Laboratory for Clinical Genomics and Advanced Technology in the Department of Pathology and Laboratory Medicine of the Dartmouth Hitchcock Health System.

## Author contributions statement

G.J.T. and W.A.K contributed to study concept and design. R.E.B and W.A.K. contributed to experiment set-up, data acquisition, and analysis. All authors contributed to interpretation of data and critical review of the manuscript.

## Competing Interests

The authors declare no competing interests.

## Additional information

### Supplementary information

None

**Correspondence** and requests for material should be addressed to G.J.T.

## Notes

### Competing Interest Statement

The authors have declared no competing interest.

### Funding Statement

no external funding was received

### Author Declarations

The Institutional Review Board at Dartmouth-Hitchcock Medical Center, has determined that this project is not research involving human subjects as defined by DHHS and FDA regulations. The study identification number that was submitted to our IRB for approval is STUDY02000805 and is considered exempt; therefore further IRB review is not required. A letter to this effect was also provided by our IRB and will be emailed to the Journal at

